# Exploring the discrepancies between clinical trials and real-world data by accounting for *Selection criteria, Operations,* and *Measurements of Outcome*

**DOI:** 10.1101/2024.01.22.24301594

**Authors:** Luca Marzano, Adam S. Darwich, Asaf Dan, Salomon Tendler, Rolf Lewensohn, Luigi De Petris, Jayanth Raghothama, Sebastiaan Meijer

**Affiliations:** Division of Health Informatics and Logistics, School of Engineering Sciences in Chemistry, Biotechnology and Health (CBH), KTH Royal Institute of Technology, Stockholm, Sweden; Dept. of Oncology-Pathology, Karolinska Institutet and the Thoracic Oncology Center, Karolinska University hospital, Stockholm, Sweden; Department of Medicine, Memorial Sloan Kettering Cancer Center, New York, NY

**Keywords:** clinical trials, oncology, trial simulation, survival analysis, population analysis, personalized medicine, chemotherapy, cancers, biostatistics, bioinformatics

## Abstract

The potential of real-world data to inform clinical trial design and supplement control arms has gained much interest in recent years. The most common approach relies on reproducing control arm outcomes by matching real-world patient cohorts to clinical trial baseline populations. However, recent studies pointed out that there is a lack of replicability, generalisability, and consensus. Further, few studies consider differences in operational processes. Discovering and accounting for confounders, including hidden effects related to the treatment process and clinical trial study protocol, would potentially allow for improved translation between clinical trials and real-world data. In this paper, we propose an approach that aims to explore and examine these confounders by investigating the impact of selection criteria and operations on the measurements of outcome. We tested the approach on a dataset consisting of small cell lung cancer patients receiving platinum-based chemotherapy regimens from a real-world data cohort (n=223) and six clinical trial control arms (n=1,224).

The results showed that the discrepancy between real-world and clinical trial data potentially depends on differences in both patient populations and operational conditions (*e.g.*, frequency of assessments, and censoring), for which further investigation is required.

The outcomes of this work suggest areas of improvement for systematically exploring and accounting for differences in outcomes between study cohorts. Continued development of the method presented here could pave the way for transferring learning across clinical studies and developing mutual translation between the real-world and clinical trials to inform clinical study design.

## Introduction

During the past few years discussions have highlighted the limitations of randomised clinical trials (RCTs) due to their high costs and the challenge of translating clinical outcomes between RCT cohorts and real-world patient populations^1,2^. Since the 21^st^ Century Cures Act in 2015, the potential for translating between real-world data (RWD) and clinical trials to inform regulatory decision-making has gained attention^3–7^. A growing body of research has focused on the extrapolation of RWD to inform clinical trial design, often described as emulation of RCT control arms or simulated (synthetic) data^8–12^, with the purpose of reproducing clinical trial outcomes^8–11,13–17^.

In an attempt to adjust for confounders, analyses have often focused on recreating the inclusion criteria of clinical trials in RWD cohorts^8–10,14,16,18–26^. The most common method is patient matching based on propensity score^27^. Propensity score is formally defined by Rosenbaum and Rubin as the conditional probability of assignment to a particular treatment^28^. The method gives a probability of an RWD patient being enrolled in a clinical trial study arm given a vector of observed covariates. Then, the RWD cohort is adjusted by including only patients with high propensity scores. Propensity score approaches, including variations of this method, still constitute the main proposed technique for adjusting for confounders in RWD cohorts based on the characteristics of RCT control arms^8,10,16,19,21,22,24,26,29,30^.

However, propensity score approaches have been shown to be limited in this aspect since a probabilistic empirical approach is highly sensitive to undetected confounders and biases of the data^1,8,10,11,14,25^. Indeed, alignment of patient characteristics is seldom sufficient to reproduce outcomes across populations^10,11,16^. It has repeatedly been pointed out that there is a lack of replicability and generalisability^1,9,10,13,17^ with only few clinical trials being replicable based on RWD^10,13,17^. Results have been mixed depending on the specific case study^13,17,31^ and measurements of outcome^10,16^ (*e.g.*, overall survival and intermediate endpoints^10,14^).

The lack of translatability has been attributed to the differences in populations, such as baseline confounders and key eligibility criteria that are not available in the data^10,11,13,14^. The main focus of improvement has been on how patient demographics affect measurements of outcome with little focus on the operations and processes behind the data^8,10,13,23,32^. The potential operational differences between clinical practice and clinical trial protocols have been mentioned as potential confounders but are yet to be fully explored^1,2,8,10,11,13,32^. There is a need to investigate the bias that is introduced by differences in investigation and clinical assessment^1,10,11,23,32^ (*e.g.*, lack of pre-trial monitoring in clinical practice^1^), and potential differences in the RCT monitoring process compared to real-world patients, with more detailed and potentially more frequent follow-up on tumour response and adverse effects, and decision-making such as withdrawal of therapy^1,10,11,25,32^.

Translation between clinical trials and real-world populations is therefore still not completely understood^1,8–10,13,14,16,17^. Further refinement of proposed methodologies are required to realise the potential of RWD to inform clinical trial design^6,9,10,13,33^. In the past, the added value of a mechanistic systems view of translation in drug development has been beneficial for other areas of model-informed drug development, such as quantitative *in vitro-in vivo* extrapolation and physiologically-based pharmacokinetics of metabolic drug-drug interactions^34^. Developing systems approaches for real-world evidence could enable a similar learn-confirm cycle and learning across studies, where the represented systems include not only the patient, disease and treatment, but also the operational context^35^.

In this paper we propose an approach that aims to systematically explore the discrepancies between RWD and RCTs by discovering and accounting for the differences in population samples (randomisation) and operation (protocols and clinical practice). We abbreviate the approach as SOMO as it is based on exploring the effects of Selection Criteria (S), Operations (O) and study protocols, on the replication of the Measurements of Outcome (MO). We developed and tested the SOMO approach using RCTs and RWD on extensive-disease (ED) small cell lung cancer (SCLC) patients receiving platinum etoposide chemotherapy. This was done to the extent it was possible given the available information, many factors were determined to be known unknowns.

## Methods

### The SOMO approach

Figure 1 shows the SOMO approach and the accompanying data analysis that was carried out. In short, the analysis included the following components:

**Figure 1.**
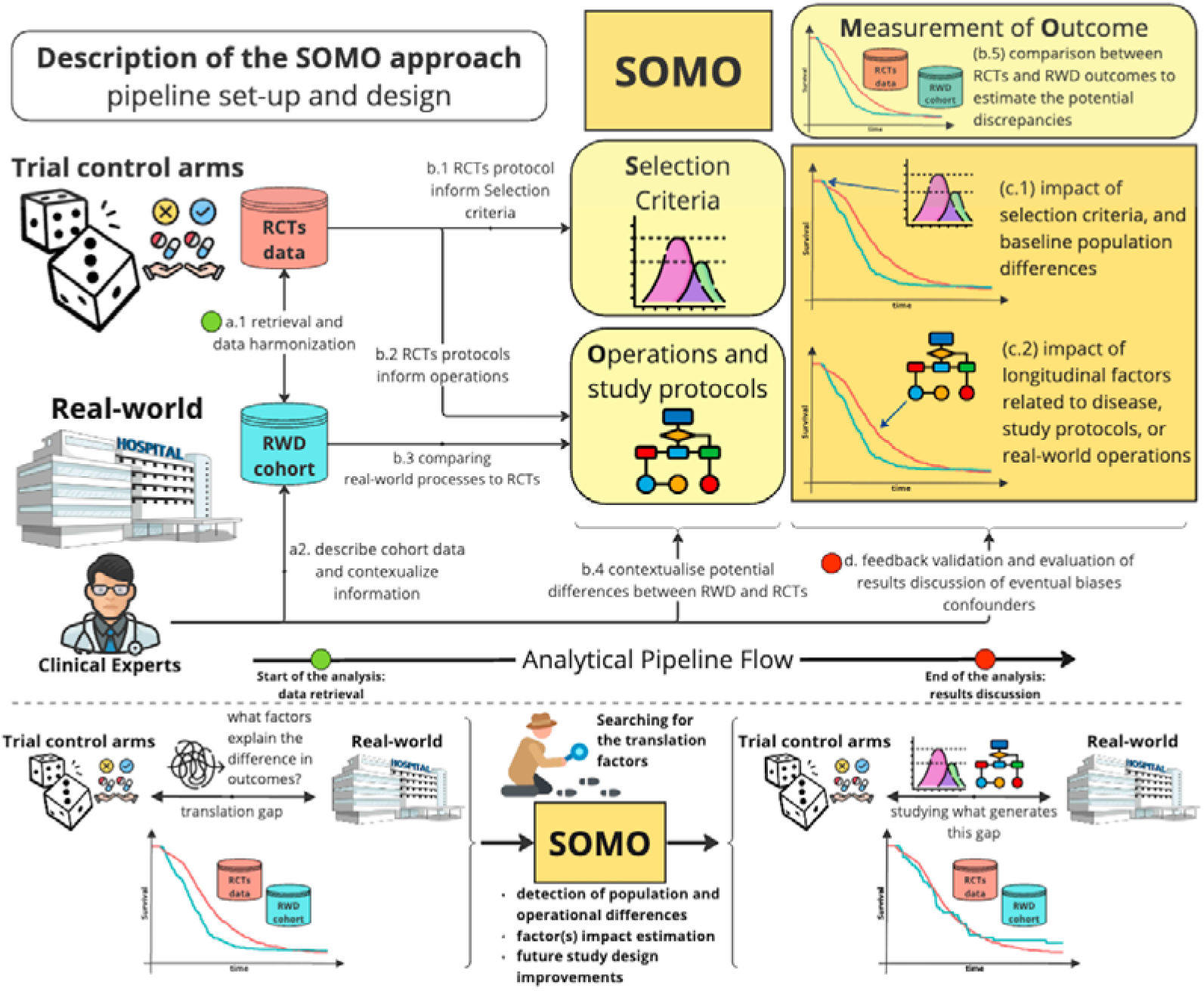
Summary description of the SOMO approach. Data are retrieved and pre-processes (step a), explorative analysis (step b), estimating the impact of factors on translation between cohorts (step c), validation and evaluation of the results (step d).

**(S). Selection Criteria** refers to all aspects related to the baseline variables that define the population, the biology of the disease, and the inclusion and exclusion criteria of the clinical datasets.

**(O). Operations and study protocols** refer to aspects related to operational processes (or mechanisms) occurring during treatment. These can be grouped into longitudinal disease factors (*e.g.*, tumour progression), the study protocol operations (*e.g.*, removal and censoring of patients that did not adhere to the study protocol), or to the potential differences from the real-world routine healthcare operations (*e.g.*, adjustment of doses, change of treatment due to relapse, patients opting to not receive treatment).

**(MO). Measurements of Outcome** refer to the metrics used to evaluate treatment efficacy (and safety, when available) and estimation of the feasibility of translation between RWD and RCTs using a comparative approach. These can be one or multiple outcomes depending on the study endpoints and the statistical analysis defined by the protocol (*e.g.*, overall survival, progress free survival, toxicity, exposure, or overall response, *etc*.), as well as and the available information from the real-world cohort.

Available RCT and RWD data are retrieved, pre-processed, and harmonised into a combined dataset. Then, clinical experts interpret and contextualise information (Figure 1, steps a.1-2). The analysis is then carried out in the following steps:

- **Explorative analysis.** An exploratory data analysis is performed to define the SOMO components (steps b.1-5). During this phase outcomes, relevant available factors, and potentially important missing aspects, are mapped into the three categories as detailed below. First, the comparison between RCTs and RWD outcomes is carried out to estimate baseline differences in measurements of outcome between cohorts. Then, a comparison between the two populations is performed to detect any potential mismatch between patient covariates due to selection and randomisation. Finally, for the operational aspects, potential confounders are investigated through comparison between study protocols and clinical practice with the aid of longitudinal outcome measures (*e.g.*, Kaplan Meier or dose response curves) and clinical expert feedback.
- **Estimating the impact of factors on the discrepancies in outcomes between cohorts.** The potential impact of selection criteria and operations is explored by applying matching based on the relevant available variables and simulating effects of operations. The impact on measurements of outcome is then compared (steps c.1-2). Clinical expert involvement allows verification of the results by identifying and discussing eventual biases and confounders (step d).

Hence, one of the outcomes was a list of potential factors related to selection criteria and operations that could contribute to explaining the translational gap and quantifying their effects, or indicative future aspects to explore when the hypotheses cannot be tested due to the lack of available information.

Case study: extensive disease small cell lung cancer patients receiving platinum-etoposide chemotherapy.

### Cohort description

In this study a mixed cohort was collated, including RWD and RCTs of ED-SCLC patients that had received platinum etoposide chemotherapy as first line treatment:

- The RWD included in this analysis was part of a retrospective cohort of SCLC patients treated at Karolinska University Hospital (Stockholm, Sweden) between 2008 and 2016^36^ (RWD KI, n patients=223). The study was approved by the institutional review boards at Karolinska Institutet and Stockholm County Council (2016/8-31).
- The RCT comparative groups originated from open data shared through the Project Data Sphere Initiative^37^, including participants receiving the standard platinum etoposide treatment that were randomised into the control arm from three randomised phase III clinical trials: PDS_Amgen (NCT00119613, n=232), PDS_Alliance (NCT00003299, n=270), PDS_EliLilly (NCT00363415, n=370), and three phase Ib-II trials PDS_PHASE2_Alliance (NCT00453154, n=46), PDS_PHASE2_EliLilly (NCT01439568, n=41), PDS_PHASE2_G1Thera (NCT02499770, n=37). A subset of patients (n=85) of PDS_EliLilly were censored after the study was declared futile after the interim analysis. These were removed prior to the analysis.

The combined mixed cohort encompassed n=1,224 patients in total. The common patient variables were age, sex, brain metastasis (BM), Eastern Cooperative Oncology Group performance status (ECOG), the cohort from which the data were retrieved (STUDY), and the label referring to if patients originated from the RCTs or real-world cohort (STUDY_TYPE), progression-free survival (PFS), Overall Survival (OS), and eventual censoring (CENSOR). The real-world cohort had been re-staged using the 8^th^ version of the International Association for the Study of Lung Cancer (IASLC) TNM in a previous validation study^38^. A stage variable (STAGE) was created where real-world patients were defined using TNM staging (IVA or IVB), while the random clinical trials patients were staged using the traditional Veterans’ Administration Lung Study Group (VALSG) method (ED stage). A summary of the real-world data and clinical trials cohorts is reported in the Supplementary Material, Table S1.

### Survival Analysis

Table 1 summarises how the SOMO approach was applied to the ED-SCLC case study. The main techniques used during the analysis were: cohort stratification given the available variables, propensity score matching with weights computed using logistic regression^27^, and oversampling to generate simulated cohorts using the standard Synthetic Minority Over-sampling TEchnique-Nominal Continuous (SMOTENC) algorithm based on k-nearest neighbours^39^. Table 1 details how these techniques were used for investigating the impact of variables on the observed outcomes.

**Table 1.**
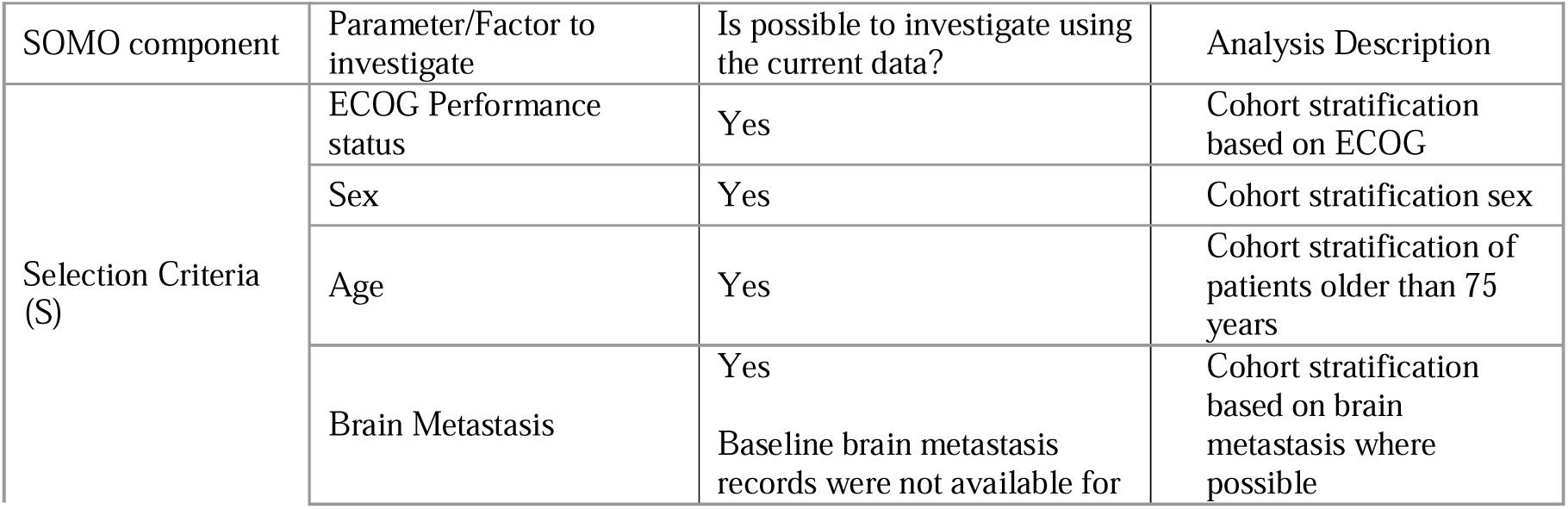
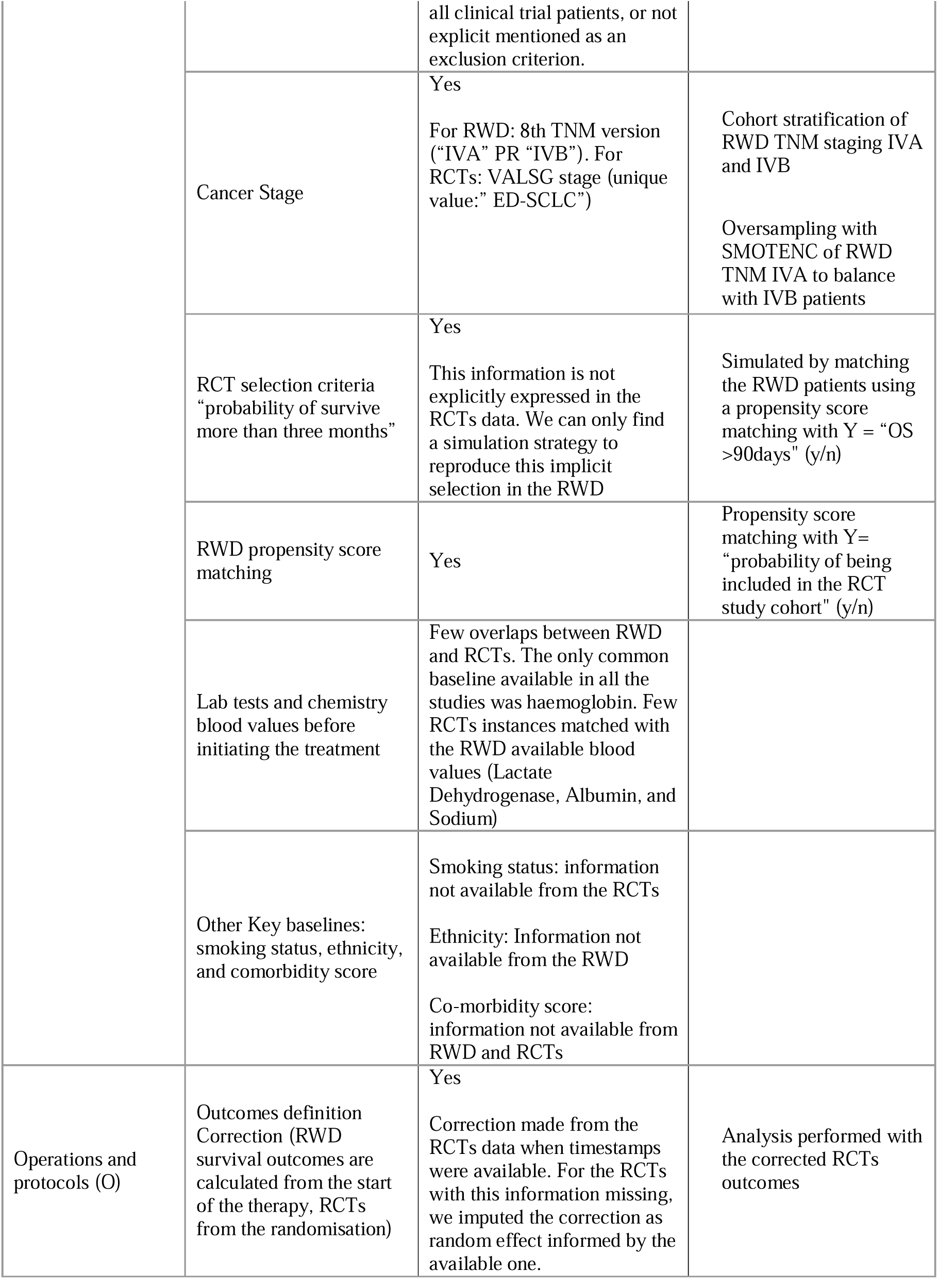
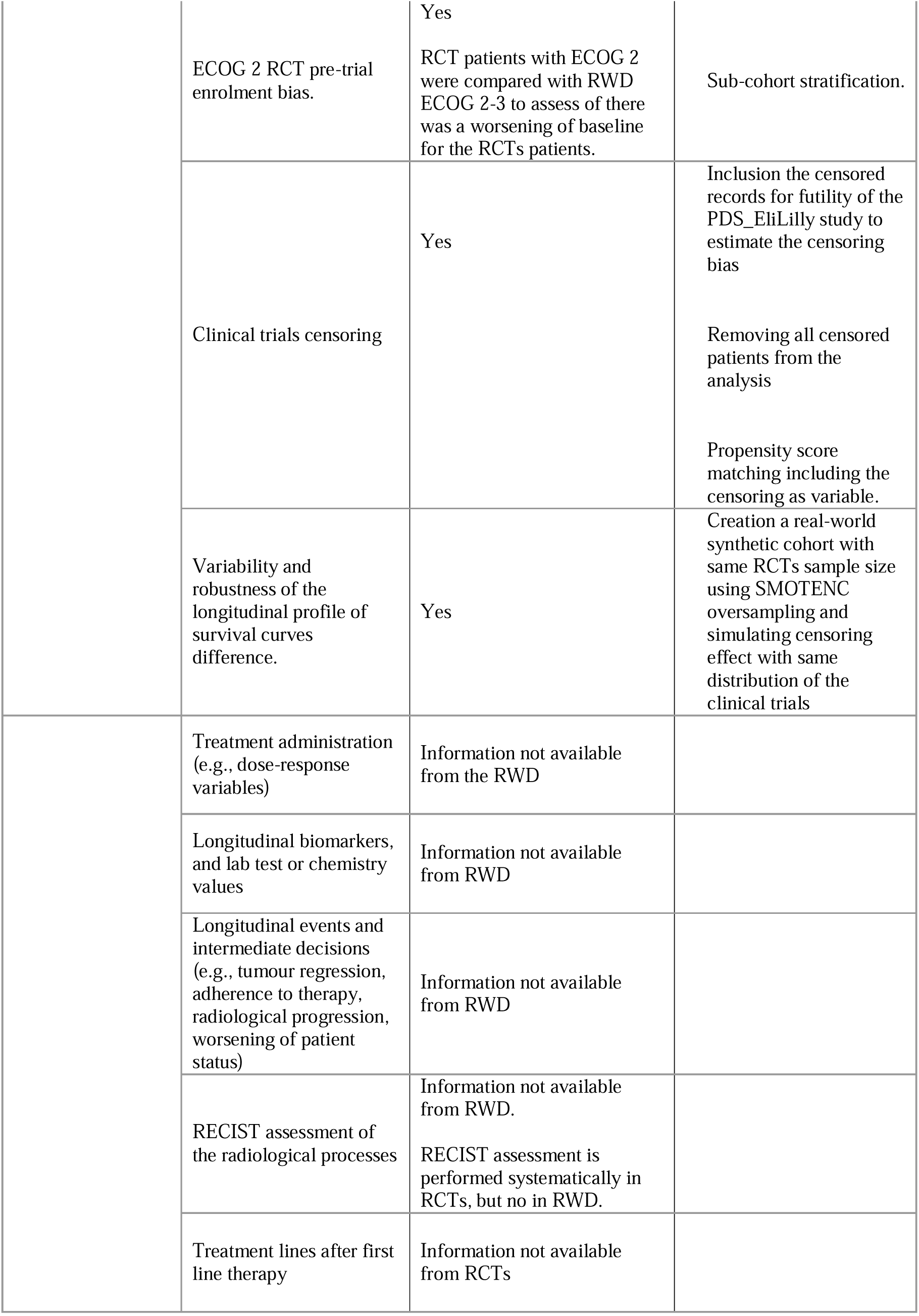
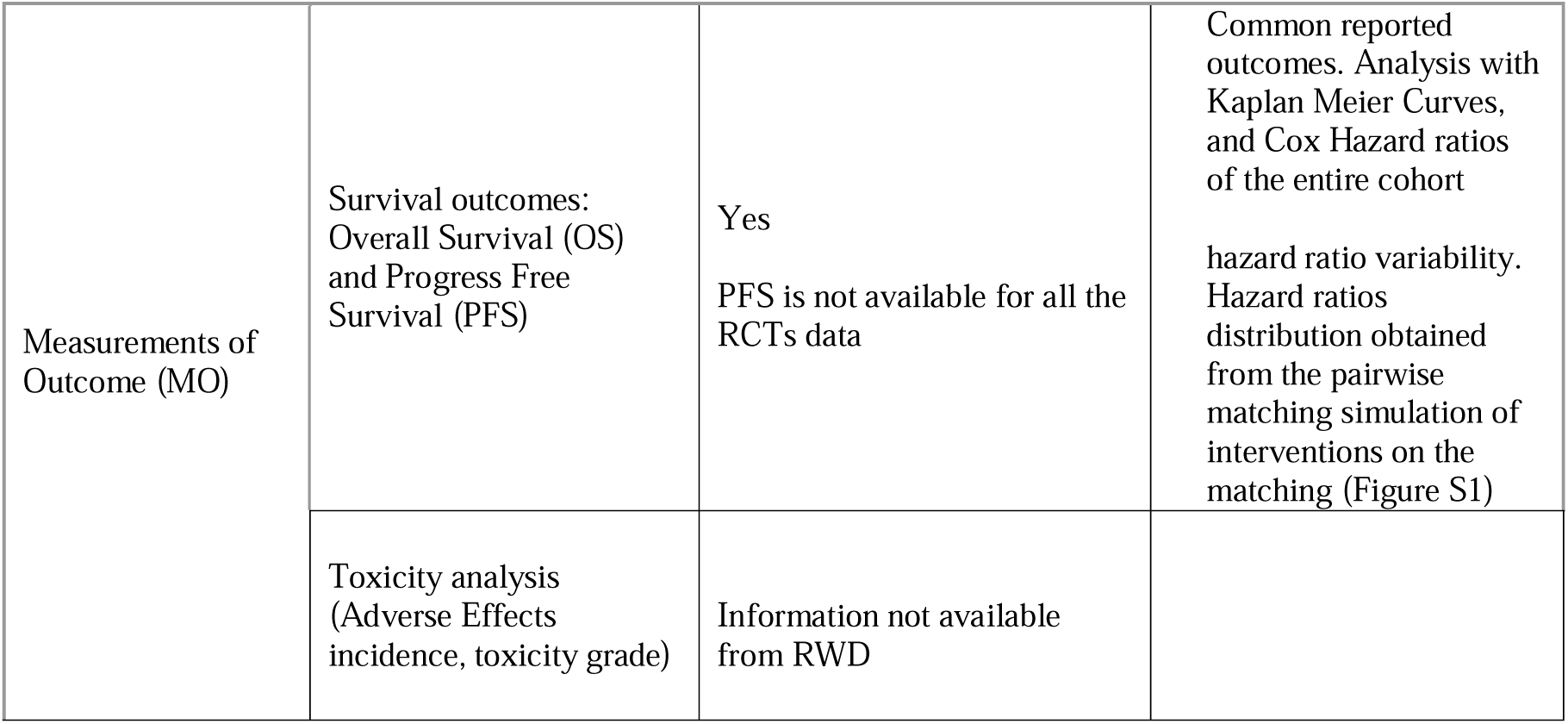
SOMO analysis description for mixed cohort SCLC study. RWD: real-world data, RCT: random clinical trial, RECIST: Response evaluation criteria in solid tumors, SMOTENC: Synthetic Minority Over-sampling TEchnique-Nominal Continuous, OS: overall survival, PFS: progress free survival.

| SOMO component | Parameter/Factor to investigate | Is possible to investigate using the current data? | Analysis Description |
| --- | --- | --- | --- |
| Selection Criteria (S) | ECOG Performance status | Yes | Cohort stratification based on ECOG |
|  | Sex | Yes | Cohort stratification sex |
|  | Age | Yes | Cohort stratification of patients older than 75 years |
|  | Brain Metastasis | Yes<br>Baseline brain metastasis records were not available for all clinical trial patients, or not explicit mentioned as an exclusion criterion. | Cohort stratification based on brain metastasis where possible |
|  | Cancer Stage | Yes<br>For RWD: 8th TNM version ("IVA" PR "IVB"). For RCTs: VALSG stage (unique value:" ED-SCLC") | Cohort stratification of RWD TNM staging IVA and IVB<br>Oversampling with SMOTENC of RWD TNM IVA to balance with IVB patients |
|  | RCT selection criteria "probability of survive more than three months" | Yes<br>This information is not explicitly expressed in the RCTs data. We can only find a simulation strategy to reproduce this implicit selection in the RWD | Simulated by matching the RWD patients using a propensity score matching with Y = "OS >90days" (y/n) |
|  | RWD propensity score matching | Yes | Propensity score matching with Y = "probability of being included in the RCT study cohort" (y/n) |
|  | Lab tests and chemistry blood values before initiating the treatment | Few overlaps between RWD and RCTs. The only common baseline available in all the studies was haemoglobin. Few RCTs instances matched with the RWD available blood values (Lactate Dehydrogenase, Albumin, and Sodium) |  |
|  | Other Key baselines: smoking status, ethnicity, and comorbidity score | Smoking status: information not available from the RCTs<br>Ethnicity: Information not available from the RWD<br>Co-morbidity score: information not available from RWD and RCTs |  |
| Operations and protocols (O) | Outcomes definition Correction (RWD survival outcomes are calculated from the start of the therapy, RCTs from the randomisation) | Yes<br>Correction made from the RCTs data when timestamps were available. For the RCTs with this information missing, we imputed the correction as random effect informed by the available one. | Analysis performed with the corrected RCTs outcomes |
|  | ECOG 2 RCT pre-trial enrolment bias. | Yes<br>RCT patients with ECOG 2 were compared with RWD ECOG 2-3 to assess if there was a worsening of baseline for the RCTs patients. | Sub-cohort stratification. |
|  | Clinical trials censoring | Yes | Inclusion the censored records for futility of the PDS_Eli Lilly study to estimate the censoring bias<br>Removing all censored patients from the analysis<br>Propensity score matching including the censoring as variable. |
|  | Variability and robustness of the longitudinal profile of survival curves difference. | Yes | Creation a real-world synthetic cohort with same RCTs sample size using SMOTENC oversampling and simulating censoring effect with same distribution of the clinical trials |
|  | Treatment administration (e.g., dose-response variables) | Information not available from the RWD |  |
|  | Longitudinal biomarkers, and lab test or chemistry values | Information not available from RWD |  |
|  | Longitudinal events and intermediate decisions (e.g., tumour regression, adherence to therapy, radiological progression, worsening of patient status) | Information not available from RWD |  |
|  | RECIST assessment of the radiological processes | Information not available from RWD.<br>RECIST assessment is performed systematically in RCTs, but not in RWD. |  |
|  | Treatment lines after first line therapy | Information not available from RCTs |  |
| Measurements of Outcome (MO) | Survival outcomes: Overall Survival (OS) and Progress Free Survival (PFS) | Yes<br>PFS is not available for all the RCTs data | Common reported outcomes. Analysis with Kaplan Meier Curves, and Cox Hazard ratios of the entire cohort<br>hazard ratio variability. Hazard ratios distribution obtained from the pairwise matching simulation of interventions on the matching (Figure S1) |
|  | Toxicity analysis (Adverse Effects incidence, toxicity grade) | Information not available from RWD |  |

First, the RCTs were combined into one dataset and directly compared with the RWD. Then, a pairwise analysis was carried out using a simulation approach to compare the RWD with each RCT to explore the between study variability in measurements of outcome.

Differences in the survival outcomes (OS and PFS) were explored. This was done using Kaplan Meier Curves and Cox proportional Hazard ratios. The reference for computing the hazard ratios was the RWD cohort.

For the pairwise analysis, simulations were carried out creating a surrogate cohort (n=250) by randomly selecting a 1:1 ratio of patients from the two populations, the RWD and the analysed RCT (Figure S1). Then, the Cox hazard ratios between real-world and clinical trials cohorts were computed. This was repeated 100 times to estimate the variability in the outcomes. Different scenarios were simulated by adjusting the selection of patients according to the identified variables across the datasets (*e.g.*, stratification by individual variables). The main assumption was that in the ideal scenario, where we would be capable of blindly adjusting for all confounders in the RWD to RCTs, there would be no significant differences in outcome between the two populations. Hence, assessing the impact of the explored factors on measurements of outcome could be used for future trial designs.

## Results

### Mapping available data and SOMO components

In total eight selection criteria factors were identified, nine potential operational discrepancies, and three potential effects on measurements of outcome. Among these, it was possible to study the effects of seven selection criteria and four operational factors, on the two measurements of outcome (See Table 1 for more details).

From the available data, it was possible to explore the effects of the commonly available variables reported in the previous section by observing the differences in outcomes for stratified subsamples of the cohort. The RWD stage IVA patients (n=46) were oversampled with the SMOTENC algorithm to balance these against the IVB patients (n=231). Propensity score matching allowed the exploration of the effects of these variables. Moreover, propensity score was used to simulate the common selection criterion across all RCTs, patients that are expected to live more than three months, for the RWD.

The operational aspects that were possible to explore were: discrepancies in outcome definitions (RWD survival outcomes were calculated from the start of the therapy, for RCTs this was defined from the randomisation), the potential progression of ECOG from 2 to 3 in the RCTs as compared to the outcomes of ECOG 3 RWD patients, and RCT censoring effects. The censoring was studied from different perspectives: removal of all censored patients, simulating censoring in a synthetic RWD cohort using SMOTENC sampling using the same distribution in the RCTs, and including the censoring as a variable in the propensity score matching.

Due to lack of overlap in information across RWD and RCTs, it was not possible to investigate the effect of blood chemistry values and biomarkers, dose administration, intermediate decisions, and longitudinal events (tumour progression or adverse effects), and treatments after the first chemo-cycle. Considerations regarding toxicity outcomes were not possible due to the lack of longitudinal information. Furthermore, information on PFS was limited and only reported for a small subset of RCT patients (mainly from PDS_EliLilly).

### Cohort discrepancies

Table 2 reports the baseline discrepancies between the studies and the main results of the comparison of RWD with the aggregated RCT data. A significant difference was observed when comparing survival outcomes in RWD and RCT patients: OS (hazard ratio: 0.65 [0.55, 0.75], reference: RWD) and PFS (hazard ratio: 0.70 [0.58, 0.85]). The full set of results are reported in the Supplementary Material, Table S2.

**Table 2.**
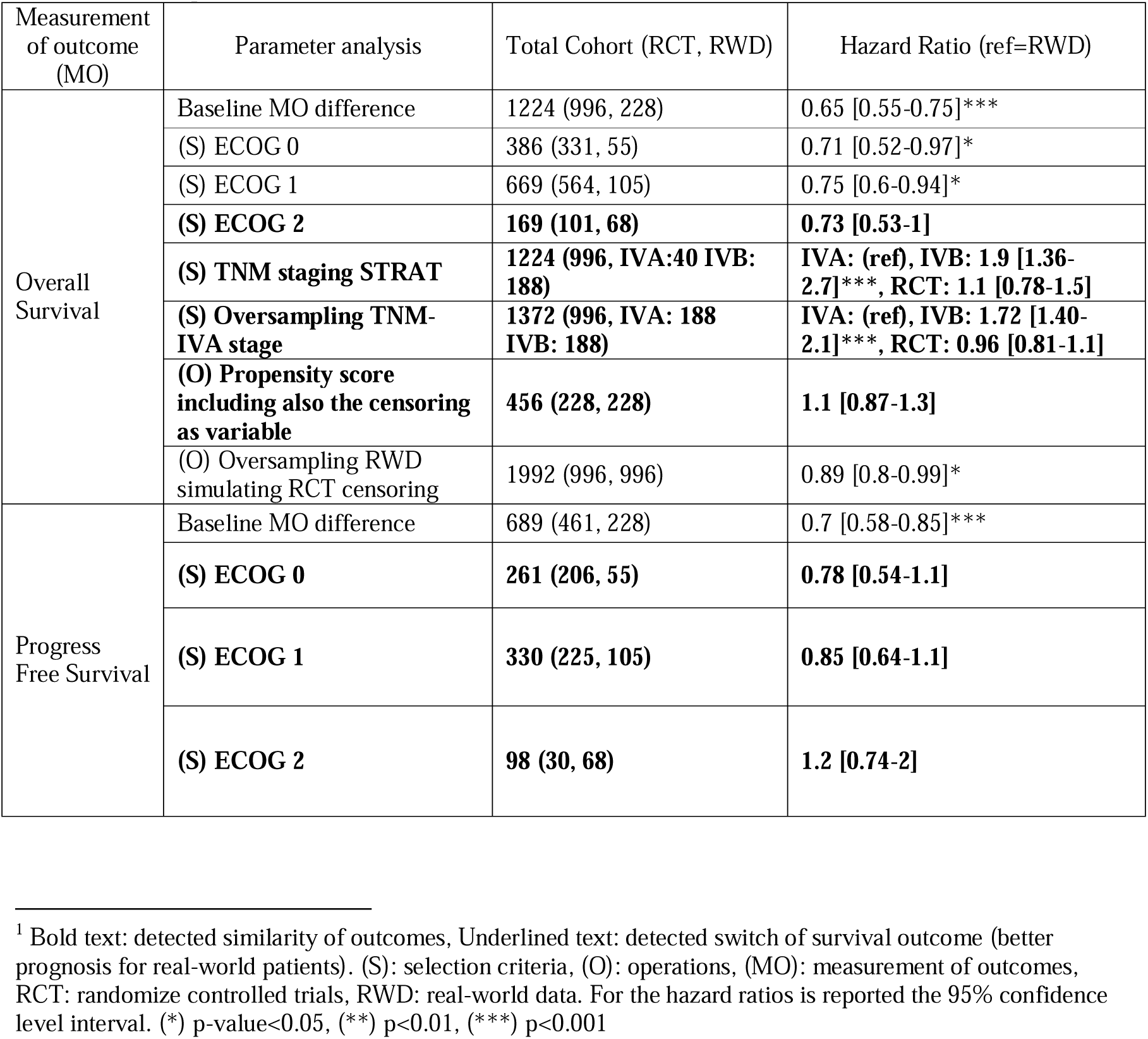
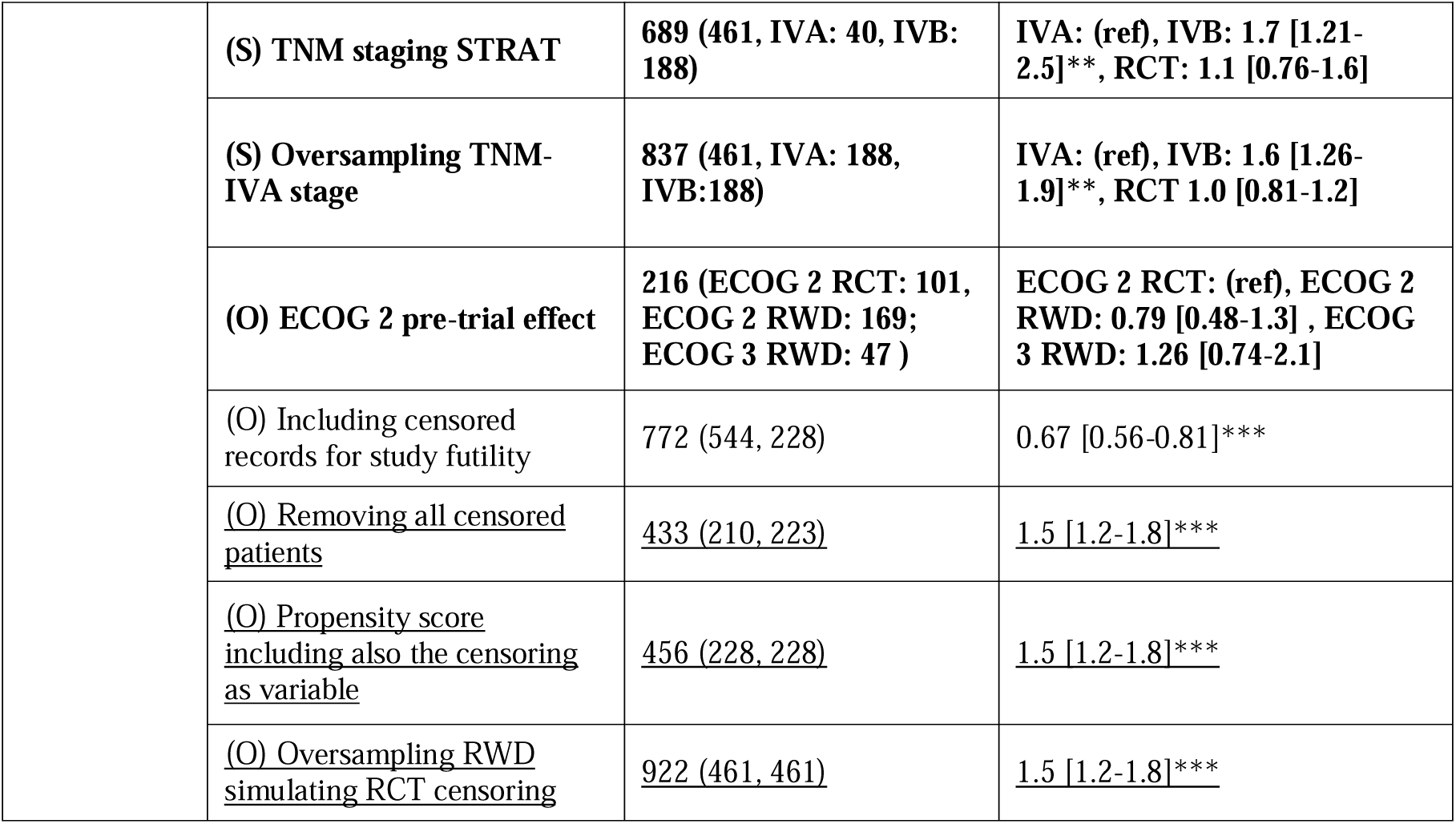
Summary of the most important results for the aggregated cohort analysis. The log-rank test of Kaplan Meier Curves confirmed the results obtained with the Cox Hazard ratios^1^.

Analysis of selection criteria identified several differences in the RWD as compared to the RCTs. In the RWD, the age distribution was skewed towards older age (median: 70 years, range: [42-86] years), a more balanced sex ratio (male: 43.4%, female: 56.6%), and a higher frequency of patients with ECOG 2 (n=68, 29.8%; Table S1). Baseline brain metastasis records were not available for all clinical trial patients, or not explicit mentioned as an exclusion criterion (*e.g.*, PDS_Alliance).

The analysis of operational aspects highlighted differences in the censoring between the two settings. Real-world censoring corresponded to a few patients (n=5) with long survival (subject to right censoring). For trial patients censoring occurred with higher frequency (i.e., n=225, 60.8% in PDS_EliLilly) across the survival range of 0-300 days. This resulted in higher estimates of OS in the RCTs compared to the RWD (Figure 2a).

**Figure 2.**
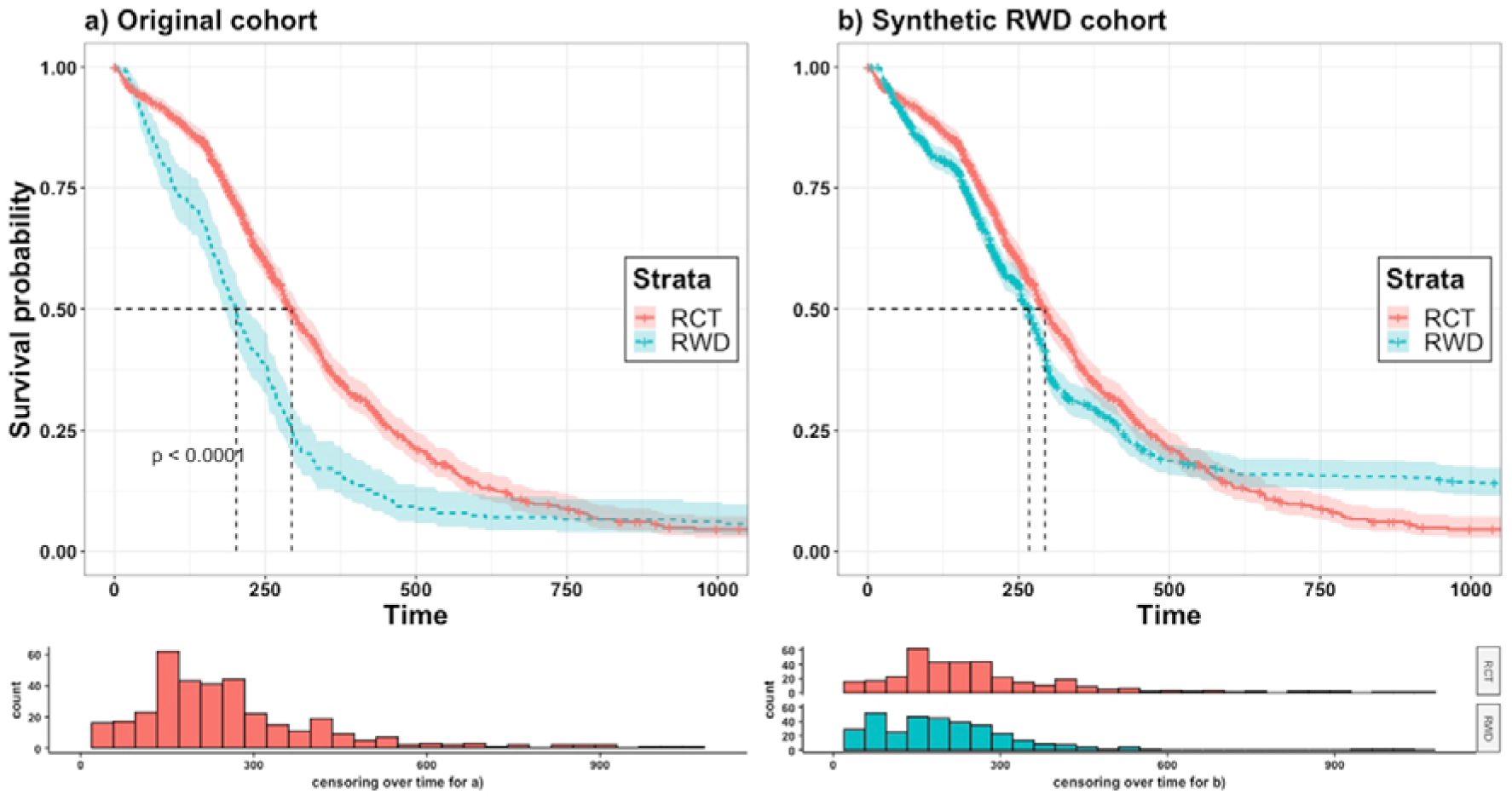
Overall Survival Kaplan Meier Curves and number of censored patients across the time for the a) baseline survival gap of measurement of outcome (n=1224), b) synthetic oversampled RWD cohort with same trial censoring (n=1992). The cohort in b) was obtained by oversampling RWD with the SMOTENC algorithm to match RCT sample size, and simulating RWD censoring using the same distribution of the RCT cohort. RWD: real-world data.

### Selection Criteria: Performance Status and TNM Staging

The subcohorts of patients that showed similar OS across the two cohorts (i.e., hazard ratios not statistically different from 1) were ECOG 2 patients (Table 2), and RWD patients with TNM staging IVA, using both conventional stratification and SMOTENC (Table 2 and Figure S2). Traditional propensity score matching did not overcome the difference in OS between the two cohorts (hazard ratio: 0.55 [0.44, 0.70]; Table S1).

For what concerns PFS, the outcomes were similar for the RCTs and RWD cohort across ECOG subgroups, TNM IVA cancer staging of RWD patients using conventional stratification and with SMOTENC oversampling, and traditional propensity score matching (Table 2).

### Operations: the effect of RCT censoring on survival estimation and propensity score matching

The effect of operational aspects could be deduced from the comparison of the survival curve shapes. Figure 2a shows that the differing shapes of the Kaplan Meier curve between the two cohorts (*i.e.*, lower survival in the first 30 days for the RCTs, followed by a higher survival for RCTs until 130 days before reaching a similar trend for the rest of the longitudinal curve).

The high RCT censoring was a key operational aspect that was impacting on the survival discrepancies. In fact, the higher survival in the RCTs during the first 130 days was strongly related to the censoring (Figure 2a), where removing the censored patients or simulating the same censoring distribution in the RWD synthetic cohort reduced the difference in overall survival (Figure 2b and Table S1).

Moreover, adjusting the propensity score by matching the censoring operations allowed to reduce the discrepancy in OS between RCTs and RWD (Figure 3). In contrast, PFS was higher in the RWD compared to RCT cohort when correcting for censoring (hazard ratio: 1.5 [1.2, 1.8]) (Table 2).

**Figure 3.**
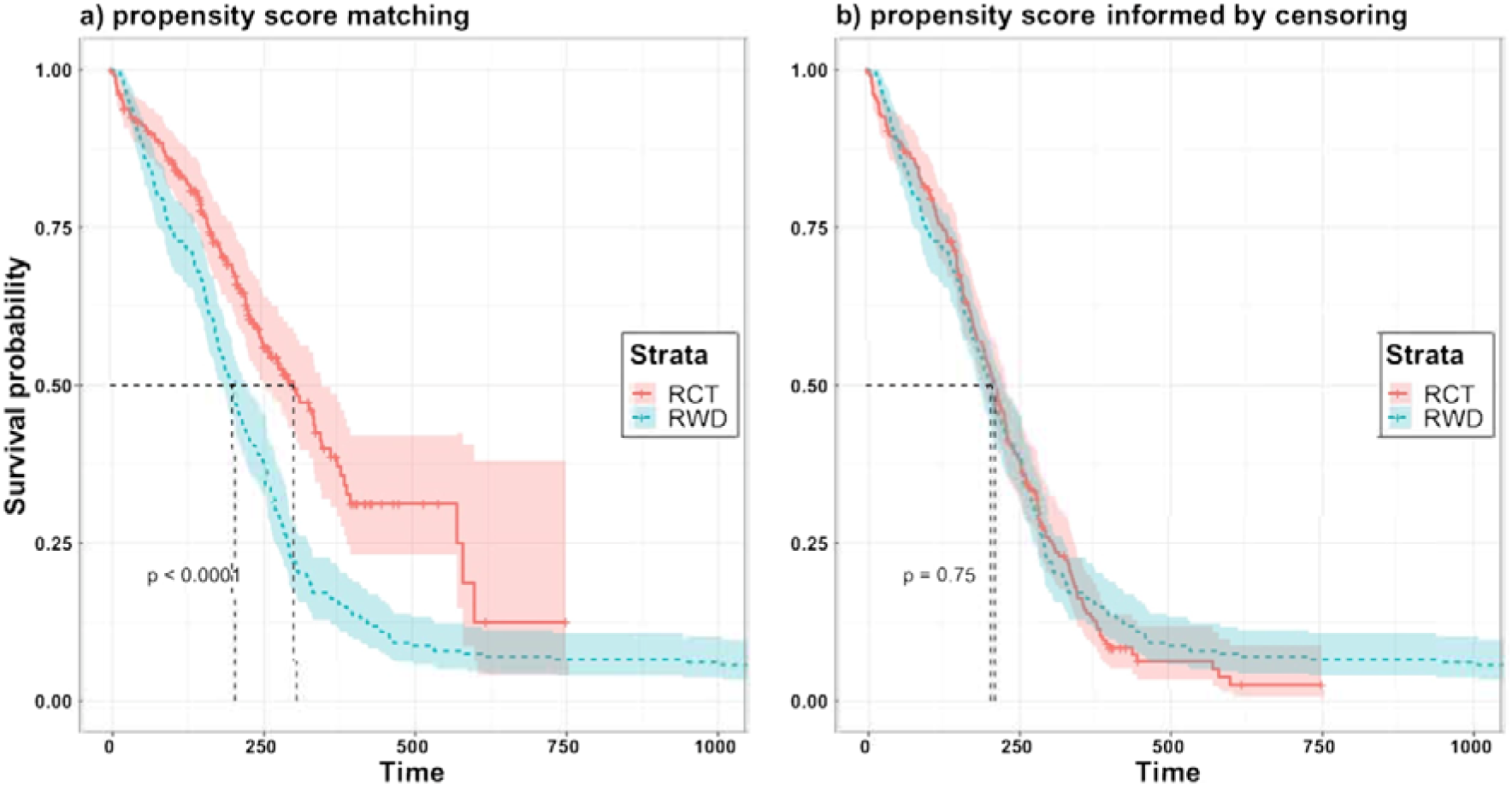
Overall Survival Kaplan Meier Curves using a) traditional propensity score marching (n=456), and b) propensity score accounting trials censoring (n=456). RWD: real-world data.

The lower OS in the RCTs during the first 30 days could be related to potential progression of baseline ECOG 2 between the time of screening and randomisation in the RCTs. In fact, Figure S3 shows that ECOG 2 patients in RCTs have similar OS as compared to ECOG 3 patients from the RWD (n=47) in the range of 0 to 30 days, and identical PFS curve.

### Pairwise randomised controlled trial comparison to the real-world population

Figures 4 and 5 show the OS of PDS_EliLilly and PDS_Amgen, and PFS for PDS_EliLilly following simulation-based oversampling of subgroups. Figure S4 shows the OS for PDS_Alliance and the aggregated cohort of Phase I-II patients following simulation-based oversampling. Overall, the results confirmed the findings of the aggregated RCT analysis (Table S2). In addition, this allowed us to investigate the variability between the RCTs and the impact of correcting for the known discrepancies between the RCTs and RWD.

**Figure 4.**
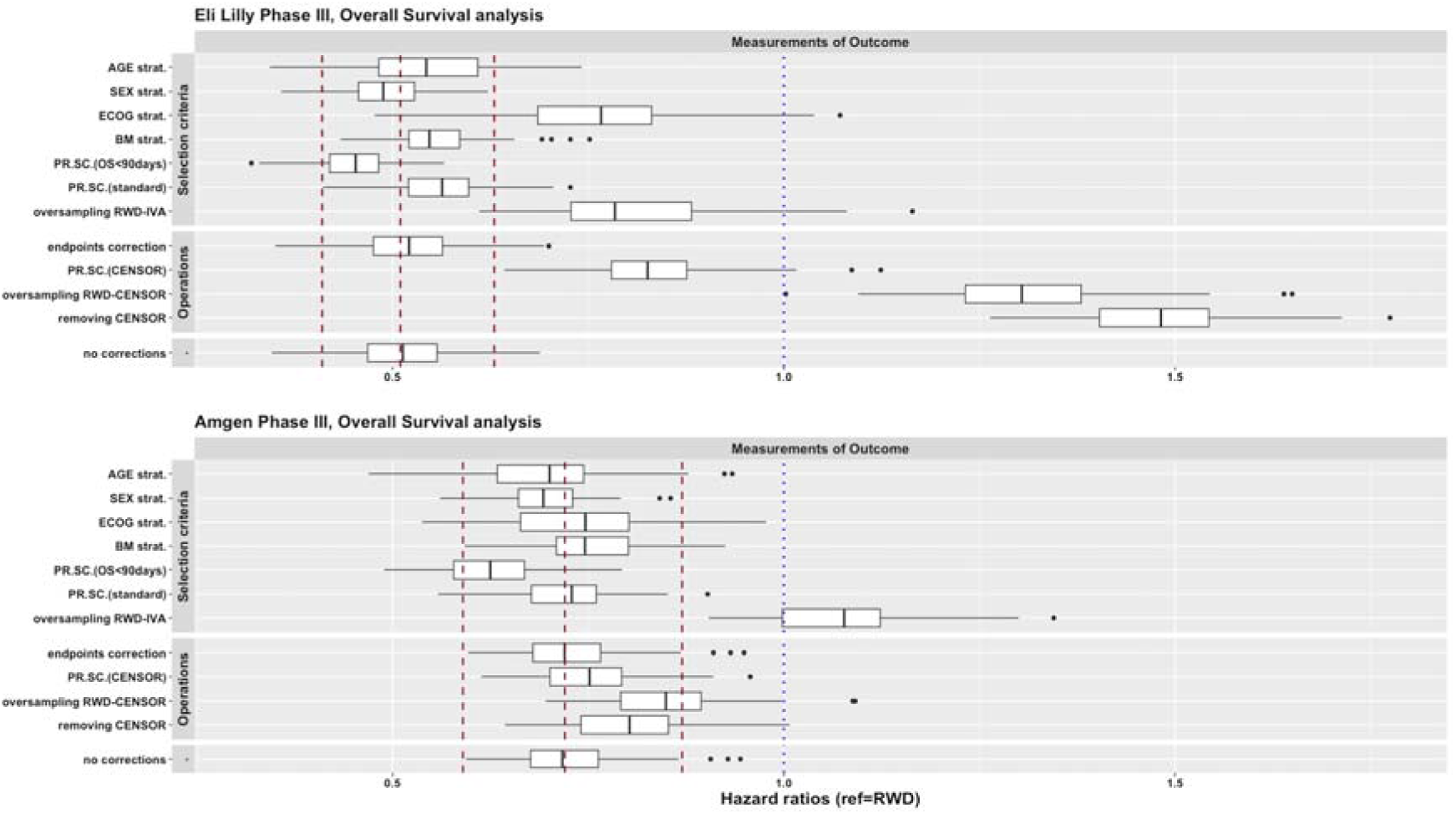
Matching Simulation results for overall survival hazard ratios for PDS_EliLilly and PDS_Amgen. Strat.: stratification, BM: brain metastases, PR.SC.: propensity score, RWD: real-world data. Hazard ratio and 95% confidence level interval with the whole cohort is reported in red dotted lines.

**Figure 5.**
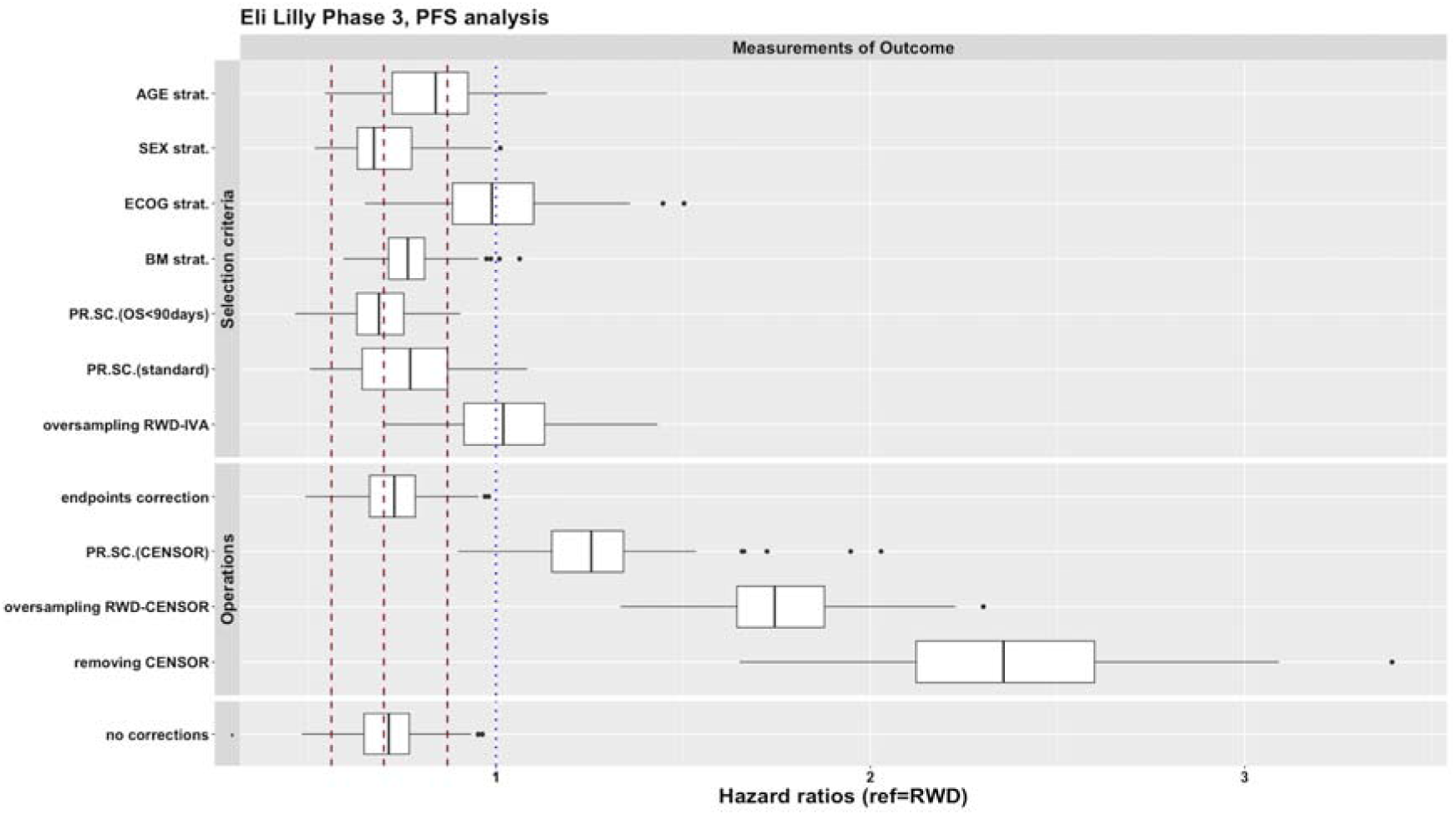
Matching Simulation results for progress free survival hazard ratios for PDS_EliLilly. Strat.: stratification, BM: brain metastases, PR.SC.: propensity score, RWD: real-world data. Hazard ratio and 95% confidence level interval with the whole cohort is reported in red dotted lines.

Oversampling of stage IVA patients in RWD played a role in reducing the discrepancies in OS across all the trials. Similarly, this improved the discrepancy in PFS for the PDS_EliLilly trial. For the trials with a relevant number of censored patients (i.e., 60% for PDS_EliLilly, 18.28% for the Phase I/II aggregated cohort), censoring was the most impactful factor and was associated with a larger difference in OS at baseline.

Figure 5 underlines the impact of operations on PFS, from which similar outcomes were achieved when correcting for ECOG and TNM stage. Instead, when correcting for censoring alone the discrepancy in PFS between PDS_EliLilly and the RWD was increased, with a. higher PFS in the RWD cohort.

## Discussion

In this study, we developed and tested an approach that aims to explore and estimate the impact of selection criteria and operations on outcome replication between RWD and RCTs. This was done using a systematic approach, attempting to account for known differences in the population samples.

Previous real-world evidence studies of lung cancer have mainly focused on non-small cell lung cancer, thus leaving small cell lung cancer understudied^40^. To the best of the authors’ knowledge, this is the most comprehensive real-world evidence study for small cell lung cancer disease. Analyses of selection criteria and measurements of outcome showed comparable results to previous studies, where a larger difference was observed in overall survival between RCTs and RWD compared to other intermediate endpoints^10,14^. Further, population differences were observed in age, an under-representation of ECOG 2 and higher representation of females in the RWD as compared to the clinical trials^41–43^.

The similarity between the stage IVA real-world patients and the clinical trial patients presented an interesting aspect. The 8^th^ TNM staging (with sub-categorisation of ED-SCLC patients in IVA and IVB stages) was not yet developed when the trials were executed, and the LD/ED staging is still largely used to define treatment intention^38^. Previous studies have pointed out significant differences in survival between IVA and IVB patients^38,44–46^. This result may be due to selection bias in RCTs, favouring younger patients with comparably better overall health and lower disease burden, which is only partially captured by the TNM staging. However, the low sample size of IVA patients in the real-world cohort is a source of potential bias and further investigation would be needed to confirm this.

The results of our work are also relevant from the general perspectives on how to improve future clinical trial design. For example, censoring in clinical trials is a known potential confounder^47^. This work underlined that this was a key operational factor with a relatively high impact, thus leading to a potential overestimation of overall survival in clinical trials (Table 2, Figure 2-3). Moreover, when censored patients were removed, other variables reported in Table S2 reduced the difference in OS between the RCTs and RWD. Thus, indicating how the censoring could bias the estimated difference in OS between the cohorts. Moreover, the interventions we applied to assess censoring effect were useful as a retrospective correction of the outcomes, but not applicable as prospective prior RWD-RCT translation.

As shown in Figure S3, ECOG 2 RCT patients showed more similar OS and PFS to ECOG 3 RWD patients. This could potentially be explained by differences in patient condition between the pre-trial phase and randomisation that could lead to worsening in baseline ECOG.

The results showed that the challenge of replicating outcomes in clinical trials from RWD patients depend not only on patient characteristics discrepancies, but also on differences in operations. This can be seen in Figures 2, 5 and S3 where the survival curves of the RCTs and RWD patients present differences in OS, potential due to operational differences in follow-up. This is underlined by the PFS analysis, where matching of population variables (performance status and TNM staging) resulted in indistinguishable PFS between RCTs and RWD. However, the discrepancy in PFS war larger when accounting for the censoring (Table 2 and Figure 5). This result is surprising, and we suspect it could be due to more frequent monitoring of clinical trial patients that could result in a shorter reported progression-free survival when adverse events or relapse occur.

In previous works, the dominant approach has been the propensity score informed by clinical trial data^10,24–26,30,31,46^. No in-depth examination of differences in operations has been done before. Figure 3 demonstrates the relevant contribution operations (i.e., censoring) to biasing propensity score matching. This can potentially explain why replication was not achieved in previous works. Indeed, in the ideal condition of having two populations with the same support of baseline covariates and sufficient population sample, accounting for the eventual operational differences, should theoretically account for any misalignment in study outcomes ^9,11,31,46^. One limitation of the propensity score is that it works from only one direction by selecting RWD patients that match RCTs, thus not accounting for any additional findings and confounders that are only detectable in the RWD population. This approach is limited for prospective applications since it would blindly work only in the scenario where all relevant confounders are accounted for in the RCTs.

RWD can inform additional disparities between RCTs and clinical practice^1^. Previous studies indicate that this difference is consistent and independent of the studied case^9,9,10,13,17^. One of the observations in this study related to the time-dynamic differences in Kaplan Meier curves between RCTs and RWD (Figure 2). Similar Kaplan Meier curves as in Figure 2a have previously been found in other oncology case studies^10,14^, thus suggesting that the impact of operations is not isolated only to small cell lung cancer. Further research is needed to understand the origins of this difference^35^. It has been noted that in translating between RCTs and RWD, understanding the complexity of clinical practice and treatment processes may be instrumental to explaining this^35^.

The SOMO approach aims to contribute to the research direction on establishing an analytical process to estimate the challenging trade-off between internal validity and generalizability^6,11,13,33^. In this work we proposed how such a framework could be beneficial for both translational directions: clinical trials can be improved by understanding the discrepancy with the real-world, and real-world therapies can be leveraged by comparing the discrepancies of trials from the real-world (*e.g.*, Figure 4 and Figure S4).

We are not claiming to have developed a definitive approach, as this work shows we identified several factors that were not possible to account for due to the lack of overlapping information. However, we believe that further development of the general method will allow for learning across translational activities. This may contribute to building regulatory acceptance in the real-world evidence approach over time as has been the case with other mechanistic modelling efforts (*e.g.*, metabolic drug-drug interaction predictions^34,48^). This is a first attempt at trying to push the real-world evidence paradigm into a similar learn-confirm cycle from which we learn case by case and pragmatically improve the usage of real-world data in future clinical trials.

The work presents some limitations to be address in the future research. There are still a set of key hypotheses reported in Table 1 that were not possible to address in the present study. The RWD represented only a single centre and lacked longitudinal variables (e.g., tumour progression, doses, and adverse effects). To address these issues, the collection of these variables and the extensive collection of data is being performed. Expansion of RWD patient cohort trough national cancer registries is the next step of the analysis. Moreover, SOMO was tested on small cell lung cancer, and research on other case studies would be beneficial to increase the generalizability of the work. In this study, the experts involved represented the clinical side of the real-world domain. In the future, we will explore the involvement of experts on clinical trial in oncology and regulatory agencies.

The increasing presence of RWD in clinical trial studies constitute a natural step for regulatory decision and future study designs. Improving upon approaches such as SOMO would pave the way to understand how we can use real-world data to close the gap between internal validity and generalizability of clinical trials.

### Study Highlights

#### What is the current knowledge on the topic?

Real-world data have the potential to inform clinical trial design, control arms, and regulatory assessment. However, real-world evidence studies have shown poor replication and generalizability and a lack of consensus on the analytical process, thus underlining that the mechanisms that would allow the translation between clinical trials and real-world populations are still not completely understood.

#### What question did this study address?

What are the mechanisms that would allow translation between clinical trials and real-world? How can we design a comprehensive and systematic approach to explore the grade of translation? Does the approach work in a challenging real-world case study such as small cell lung cancer chemotherapy?

#### What does this study add to our knowledge?

Differences in operations and protocols have a relevant impact on the gap in clinical outcomes. These must be studied in concomitance with the selection criteria of the baselines. Previous works proposed pure empirical approaches (such as propensity score), and limitations of the findings can be related to the lack of consideration of operational differences between trials and real-world practice. Our approach allowed novel insights regarding which aspects would benefit from further investigation to improve the design of small cell lung cancer studies (ECOG 2 underrepresentation and pre-trial biases, exploring the therapies with the new TNM staging categories, operational biases of trials censoring and progress free survival)

#### How might this change clinical pharmacology or translational science?

Designing a comprehensive and systematic approach to investigate how selection criteria and operations are impacting on the measurements of outcome would allow us to estimate the trade-off between internal validity and generalizability of clinical trials. Thus, pushing real-world evidence towards a learn and confirm cycle from which we learn case by case and close the translational gap between clinical trials and real-world populations.

## Data Availability

All data produced in the present study are available upon reasonable request to the authors

## Acknowledgments

This project is a contribution to the Centre for Data-Driven Health (CDDH), KTH Royal Institute of Technology (https://www.kth.se/sv/cddh).

## Authors Contribution

L.M. and A.S.D. wrote the manuscript. L.M., A.S.D., A.D., S.T., J.R., and S.M. designed the research. L.M., A.S.D., S.T., A.D., R.L., and L.D.P. performed the research. L.M., A.S.D., S.T., A.D., R.L., and L.D.P. analyzed the data

## Supplementary information Titles

- Supplementary Materials for Methods section
- Supplementary Materials for Results section

## References

1. Beaulieu-Jones, B. K. et al. Examining the Use of Real-World Evidence in the Regulatory Process. Clinical Pharmacology & Therapeutics 107, 843–852 (2020).

2. Franklin, J. M. & Schneeweiss, S. When and How Can Real World Data Analyses Substitute for Randomized Controlled Trials? Clinical Pharmacology & Therapeutics 102, 924–933 (2017).

3. Schurman, B. The Framework for FDA’s Real-World Evidence Program. Applied Clinical Trials 28, 15–17 (2019).

4. Dagenais, S., Russo, L., Madsen, A., Webster, J. & Becnel, L. Use of Real-World Evidence to Drive Drug Development Strategy and Inform Clinical Trial Design. Clinical Pharmacology & Therapeutics 111, 77–89 (2022).

5. Burns, L. et al. Real-World Evidence for Regulatory Decision-Making: Guidance From Around the World. Clinical Therapeutics 44, 420–437 (2022).

6. Baumfeld Andre, E., et al. The Current Landscape and Emerging Applications for Real-World Data in Diagnostics and Clinical Decision Support and its Impact on Regulatory Decision Making. Clinical Pharmacology & Therapeutics 112, 1172– 1182 (2022).

7. Campbell, U. B., Honig, N. & Gatto, N. M. SURF: A Screening Tool (for Sponsors) to Evaluate Whether Using Real-World Data to Support an Effectiveness Claim in an FDA Application Has Regulatory Feasibility. Clinical Pharmacology & Therapeutics 114, 981–993 (2023).

8. Wang, C. Y. et al. Uncontrolled Extensions of Clinical Trials and the Use of External Controls—Scoping Opportunities and Methods. Clinical Pharmacology & Therapeutics 111, 187–199 (2022).

9. Wang, S. V. et al. Reproducibility of real-world evidence studies using clinical practice data to inform regulatory and coverage decisions. Nature Communications 13, 1–11 (2022).

10. Jemielita, T. et al. Replication of Oncology Randomized Trial Results using Swedish Registry Real World-Data: A Feasibility Study. Clinical Pharmacology & Therapeutics 110, 1613–1621 (2021).

11. Sreeram V. Ramagopalan, Ramagopalan, S. V., Ramagopalan, S. V., Simpson, A. & Sammon, C. J. Can real-world data really replace randomised clinical trials? BMC Medicine 18, 13–13 (2020).

12. Lin, J., Liao, R. & Gamalo-Siebers, M. Dynamic incorporation of real world evidence within the framework of adaptive design. J Biopharm Stat 32, 986–998 (2022).

13. He, Z. et al. Clinical Trial Generalizability Assessment in the Big Data Era: A Review. Clinical and Translational Science 13, 675–684 (2020).

14. Tan, K. et al. Emulating Control Arms for Cancer Clinical Trials Using External Cohorts Created From Electronic Health Record-Derived Real-World Data. Clinical Pharmacology & Therapeutics 111, 168–178 (2022).

15. Franklin, J. M., Glynn, R. J., Martin, D. & Schneeweiss, S. Evaluating the Use of Nonrandomized Real-World Data Analyses for Regulatory Decision Making. Clinical Pharmacology & Therapeutics 105, 867–877 (2019).

16. Franklin, J. M. et al. Nonrandomized Real-World Evidence to Support Regulatory Decision Making: Process for a Randomized Trial Replication Project. Clinical pharmacology and therapeutics 107, 817–826 (2020).

17. Bartlett, V., Dhruva, S., Shah, N., Ryan, P. & Ross, J. Feasibility of Using Real-World Data to Replicate Clinical Trial Evidence. JAMA NETWORK OPEN 2, (2019).

18. Abrahami, D. et al. Use of Real-World Data to Emulate a Clinical Trial and Support Regulatory Decision Making: Assessing the Impact of Temporality, Comparator Choice, and Method of Adjustment. Clinical Pharmacology & Therapeutics 109, 452–461 (2021).

19. Ho, M. et al. Examples of Applying RWE Causal-Inference Roadmap to Clinical Studies. STATISTICS IN BIOPHARMACEUTICAL RESEARCH (2023) doi:10.1080/19466315.2023.2177333.

20. Stewart, M. et al. An Exploratory Analysis of Real-World End Points for Assessing Outcomes Among Immunotherapy-Treated Patients With Advanced Non–Small-Cell Lung Cancer. JCO Clinical Cancer Informatics 1–15 (2019) doi:10.1200/cci.18.00155.

21. Oksen, D. et al. Treatment effectiveness in a rare oncology indication: Lessons from an external control cohort study. Clinical and Translational Science 15, 1990–1998 (2022).

22. Rivera, D. R. et al. The Friends of Cancer Research Real-World Data Collaboration Pilot 2.0: Methodological Recommendations from Oncology Case Studies. Clinical Pharmacology & Therapeutics 111, 283–292 (2022).

23. Cramer-van der Welle, C. M., et al. Real-world outcomes versus clinical trial results of immunotherapy in stage IV non-small cell lung cancer (NSCLC) in the Netherlands. Scientific Reports 11, 6306 (2021).

24. Lakdawalla, D. N. et al. Predicting Real-World Effectiveness of Cancer Therapies Using Overall Survival and Progression-Free Survival from Clinical Trials: Empirical Evidence for the ASCO Value Framework. Value in Health 20, 866– 875 (2017).

25. Lasiter, L. et al. Real-world Overall Survival Using Oncology Electronic Health Record Data: Friends of Cancer Research Pilot. Clinical Pharmacology & Therapeutics 111, 444–454 (2022).

26. Loureiro, H. et al. Matching by OS prognostic score to construct external controls in lung cancer clinical trials. Clinical Pharmacology & Therapeutics n/a,.

27. Webster-Clark, M. et al. Using propensity scores to estimate effects of treatment initiation decisions: State of the science. Statistics in Medicine 40, 1718–1735 (2021).

28. Rosenbaum, P. R. & Rubin, D. B. The central role of the propensity score in observational studies for causal effects. Biometrika 70, 41–55 (1983).

29. Loureiro, H., Becker, T., Bauer-Mehren, A., Ahmidi, N. & Weberpals, J. Artificial Intelligence for Prognostic Scores in Oncology: a Benchmarking Study. Frontiers in artificial intelligence 4, (2021).

30. Liu, R. et al. Evaluating eligibility criteria of oncology trials using real-world data and AI. Nature 592, 629–633 (2021).

31. Franklin, J. et al. Emulating Randomized Clinical Trials With Nonrandomized Real-World Evidence Studies First Results From the RCT DUPLICATE Initiative. CIRCULATION 143, 1002–1013 (2021).

32. Feinberg, B. A. et al. Use of Real-World Evidence to Support FDA Approval of Oncology Drugs. Value in Health 23, 1358–1365 (2020).

33. Bolislis, W. R., Fay, M. & Kühler, T. C. Use of Real-world Data for New Drug Applications and Line Extensions. Clinical Therapeutics 42, 926–938 (2020).

34. Wang, Y. et al. Model-Informed Drug Development: Current US Regulatory Practice and Future Considerations. Clinical Pharmacology & Therapeutics 105, 899–911 (2019).

35. Greenhalgh, T. & Papoutsi, C. Studying complexity in health services research: desperately seeking an overdue paradigm shift. BMC Medicine 16, 95 (2018).

36. Tendler, S. et al. Treatment patterns and survival outcomes for small-cell lung cancer patients - a Swedish single center cohort study. Acta Oncol 59, 388–394 (2020).

37. Green, A. K. et al. The Project Data Sphere Initiative: Accelerating Cancer Research by Sharing Data. The Oncologist 20, 464–e20 (2015).

38. Tendler, S. et al. Validation of the 8th TNM classification for small-cell lung cancer in a retrospective material from Sweden. Lung Cancer 120, 75–81 (2018).

39. Chawla, N. V., Bowyer, K. W., Hall, L. O. & Kegelmeyer, W. P. SMOTE: Synthetic Minority Over-sampling Technique. Journal of Artificial Intelligence Research 16, 321–357 (2002).

40. Pietanza, M. C., Byers, L. A., Minna, J. D. & Rudin, C. M. Small Cell Lung Cancer: Will Recent Progress Lead to Improved Outcomes? Clinical Cancer Research 21, 2244–2255 (2015).

41. Jaoude, J. A. et al. Performance Status Restriction in Phase III Cancer Clinical Trials. Journal of the National Comprehensive Cancer Network : JNCCN 18 10, 1322–1326 (2020).

42. Pang, H. H. et al. Enrollment Trends and Disparity Among Patients With Lung Cancer in National Clinical Trials, 1990 to 2012. JCO 34, 3992–3999 (2016).

43. Hutchins, L. F., Unger, J. M., Crowley, J. J., Coltman, C. A. & Albain, K. S. Underrepresentation of Patients 65 Years of Age or Older in Cancer-Treatment Trials. New England Journal of Medicine 341, 2061–2067 (1999).

44. Hwang, J. K. et al. Validation of the Eighth Edition TNM Lung Cancer Staging System. Journal of Thoracic Oncology 15, 649–654 (2020).

45. Marzano, L. et al. A novel analytical framework for risk stratification of real-world data using machine learning: A small cell lung cancer study. Clinical and Translational Science 00, 1–11 (2022).

46. Tan, F. et al. External validation of the eighth edition of the TNM classification for lung cancer in small cell lung cancer. Lung Cancer 170, 98–104 (2022).

47. Gilboa, S. et al. Informative censoring of surrogate end-point data in phase 3 oncology trials. Eur J Cancer 153, 190–202 (2021).

48. Shebley, M. et al. Physiologically Based Pharmacokinetic Model Qualification and Reporting Procedures for Regulatory Submissions: A Consortium Perspective. Clinical Pharmacology & Therapeutics 104, 88–110 (2018).

