## Supplementary Materials for "Exploring the discrepancies between clinical trials and real-world data by accounting for *Selection criteria, Operations,* and *Measurements of Outcome*"

*Stockholm, Sweden*

*^b^Dept. of Oncology-Pathology, Karolinska Institutet and the Thoracic Oncology Center, Karolinska University hospital, Stockholm, Sweden*

*^c^Department of Medicine, Memorial Sloan Kettering Cancer Center, New York, NY*

### **Supplementary Materials for Methods section**

Table S1. Summary of the ED-SCLC mixed cohorts^[1]^.

|  | **RCT Phase III** | | | **RCT Phase I/II** | | | **Real World Data** |
| --- | --- | --- | --- | --- | --- | --- | --- |
| **ClinicalTrials.gov ID** | **NCT00003299** | **NCT00119613** | **NCT00363415** | **NCT00453154** | **NCT01439568** | **NCT02499770** |  |
| **STUDY** | **PDS_Alliance** | **PDS_Amgen** | **PDS_ELiLilly** | **PDS_PHASE2_Allianc** | **PDS_PHASE2_EliLilly** | **PDS_PHASE2_G1Thera** | **RWD KI** |
| **Sample size** | **(N=270)** | **(N=232)** | **(N=370)** | **(N=46)** | **(N=41)** | **(N=37)** | **(N=228)** |
| **AGE** |  |  |  |  |  |  |  |
| Mean (SD) | 61.0 (9.51) | 61.3 (8.09) | 62.7 (9.65) | NA (NA) | 66.7 (8.00) | NA (NA) | 68.1 (8.94) |
| Median [Min, Max] | 61.5 [34.0, 81.0] | 62.0 [37.0, 81.0] | 63.5 [38.3, 86.2] | NA [NA, NA] | 66.1 [47.8, 82.0] | NA [NA, NA] | 70.0 [42.0, 86.0] |
| Missing | 0 (0%) | 0 (0%) | 0 (0%) | 46 (100%) | 0 (0%) | 37 (100%) | 0 (0%) |
| **SEX** |  |  |  |  |  |  |  |
| F | 121 (44.8%) | 73 (31.5%) | 104 (28.1%) | 25 (54.3%) | 25 (61.0%) | 11 (29.7%) | 129 (56.6%) |
| M | 149 (55.2%) | 159 (68.5%) | 266 (71.9%) | 21 (45.7%) | 16 (39.0%) | 26 (70.3%) | 99 (43.4%) |
| **ECOG** |  |  |  |  |  |  |  |
| 0 | 79 (29.3%) | 35 (15.1%) | 168 (45.4%) | 18 (39.1%) | 15 (36.6%) | 16 (43.2%) | 55 (24.1%) |
| 1 | 178 (65.9%) | 146 (62.9%) | 179 (48.4%) | 18 (39.1%) | 24 (58.5%) | 19 (51.4%) | 105 (46.1%) |
| 2 | 13 (4.8%) | 51 (22.0%) | 23 (6.2%) | 10 (21.7%) | 2 (4.9%) | 2 (5.4%) | 68 (29.8%) |
| **BM** |  |  |  |  |  |  |  |
| Yes | 0 (0%) | 0 (0%) | 41 (11.1%) | 0 (0%) | 0 (0%) | 0 (0%) | 39 (17.1%) |
| Missing | 270 (100%) | 232 (100%) | 329 (88.9%) | 46 (100%) | 41 (100%) | 37 (100%) | 189 (82.9%) |
| **CENSOR** |  |  |  |  |  |  |  |
| Yes | 18 (6.7%) | 33 (14.2%) | 225 (60.8%) | 9 (19.6%) | 12 (29.3%) | 9 (24.3%) | 5 (2.2%) |
| No | 252 (93.3%) | 199 (85.8%) | 145 (39.2%) | 37 (80.4%) | 29 (70.7%) | 28 (75.7%) | 223 (97.8%) |
| **PFS** |  |  |  |  |  |  |  |
| Mean (SD) | NA (NA) | NA (NA) | 135 (78.1) | 90.2 (46.9) | 164 (94.6) | 330 (167) | 248 (349) |
| Median [Min, Max] | NA [NA, NA] | NA [NA, NA] | 139 [0.986, 440] | 86.6 [41.3, 178] | 149 [10.8, 400] | 288 [84.0, 805] | 175 [6.00, 3110] |
| Missing | 270 (100%) | 232 (100%) | 0 (0%) | 33 (71.7%) | 0 (0%) | 0 (0%) | 0 (0%) |
| **OS** |  |  |  |  |  |  |  |
| Mean (SD) | 354 (268) | 287 (207) | 199 (105) | 272 (194) | 311 (177) | 341 (201) | 285 (364) |
| Median [Min, Max] | 288 [0, 1950] | 261 [2.00, 1180] | 195 [0.986, 491] | 189 [74.8, 898] | 277 [10.8, 753] | 296 [59.0, 840] | 202 [6.00, 3110] |

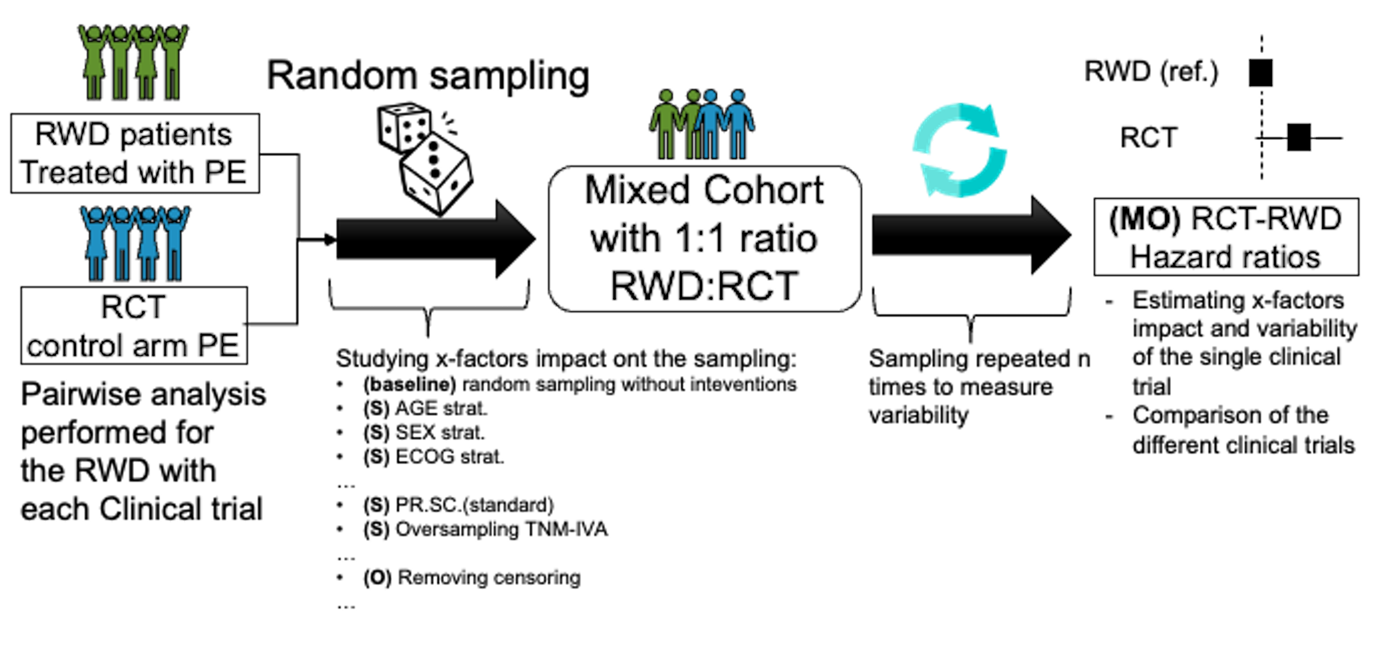

Figure S1. Bootstrapping sampling simulation to estimate impact and variability of selection criteria (S) and operations (O) impact on Cox Hazard Ratios. MO: Measurement of Outcome

### **Supplementary Materials for Results section**

Table S2. Summary of the results for the aggregated cohort analysis. The log-rank test of Kaplan Meier Curves confirmed the results obtained with the Cox Hazard ratios^[2]^.

| Measurement of outcome (MO) | Parameter analysis | Total Cohort (RCT, RWD) | Hazard Ratio (ref=RWD) |
| --- | --- | --- | --- |
| Overall Survival | Baseline MO difference | 1224 (996, 228) | 0.65 [0.55-0.75]*** |
|  | (S) ECOG 0 | 386 (331, 55) | 0.71 [0.52-0.97]* |
|  | (S) ECOG 1 | 669 (564, 105) | 0.75 [0.6-0.94]* |
|  | **(S) ECOG 2** | **169 (101, 68)** | **0.73 [0.53-1]** |
|  | (S) SEX M | 736 (637, 99) | 0.72 [0.58-0.91]** |
|  | (S) SEX F | 488 (359, 129) | 0.55 [0.44-0.68]*** |
|  | (S) AGE > 75 ys | 140 (86, 54) | 0.61 [0.42-0.9]* |
|  | (S) BM Yes | 80 (41, 39) | 0.28 [0.15-0.52]*** |
|  | (S) BM No | 1144 (955, 189) | 0.69 ]0.59-0.82]*** |
|  | **(S) TNM staging STRAT** | **1224 (996, IVA:40 IVB: 188)** | **IVA:(ref), IVB: 1.9 [1.36-2.7]***, RCT 1.1: [0.78-1.5]** |
|  | **(S) Oversampling TNM-IVA stage** | **1372 (996, IVA: 188 IVB: 188)** | **IVA:(ref), IVB: 1.72 [1.40-2.1]***, RCT 0.96: [0.81-1.1]** |
|  | (S) probability OS>three months propensity score | 226 (173, 53) | 0.12 [0.07-0.18]*** |
|  | (S) Propensity sccore matching | 456 (228, 228) | 0.55 [0.44-0.70]*** |
|  | (O) MO correction | 1224 (996, 228) | 0.65 [0.56-0.76]*** |
|  | (O) ECOG 2 pre-trial effect | 216 (101, RWD ECOG 2: 68; RWD ECOG 3: 47) | ECOG 2 RCT: (ref), ECOG 2 RWD: 1.3 [0.97-1.9], ECOG 3 RWD: 2.0 [1.37-2.8]*** |
|  | (O) Including censored records for study futility | 1336 (1108, 228) | 0.63 [0.54-0.74]*** |
|  | (O) Removing all censored patients | 913 (690, 223) | 0.85 [0.73-0.99]* |
|  | **(O) Propensity score including also the censoring as variable** | **456 (228, 228)** | **1.1 [0.87-1.3]** |
|  | (O) Oversampling RWD simulating RCT censoring | 1992 (996, 996) | 0.89 [0.8-0.99]* |
|  | Baseline MO difference | 689 (461, 228) | 0.7 [0.58-0.85]*** |
| Progress Free Survival | **(S) ECOG 0** | **261 (206, 55)** | **0.78 [0.54-1.1]** |
|  | **(S) ECOG 1** | **330 (225, 105)** | **0.85 [0.64-1.1]** |
|  | **(S) ECOG 2** | **98 (30, 68)** | **1.2 [0.74-2]** |
|  | (S) SEX M | 414 (315, 99) | 0.73 [0.56-0.96]* |
|  | (S) SEX F | 275 (146, 129) | 0.67 [0.5-0.91]* |
|  | (S) Patients > 75 ys | 105 (54, 51) | 0.59 [0.36-0.95]* |
|  | (S) BM Yes | 80 (41, 39) | 0.39 [0.2-0.74]** |
|  | (S) BM No | 609 (420, 189) | 0.76 [0.52-0.94]* |
|  | **(S) TNM staging STRAT** | **689 (461, IVA: 40, IVB: 188)** | **IVA:(ref), IVB: 1.7 [1.21-2.5]**, RCT 1.1 [0.76-1.6]** |
|  | **(S) Oversampling TNM-IVA stage** | **837 (461, IVA: 188, IVB:188)** | **IVA:(ref), IVB: 1.6 [1.26-1.9]**, RCT: 1.0 [0.81-1.2]** |
|  | (S) probability OS>three months propensity score | 358 (258, 100) | 0.54 [0.4-0.72]*** |
|  | **(S) Propensity score matching** | **456 (228, 228)** | **0.79 [0.62-1]** |
|  | (O) MO correction | 689 (461, 228) | 0.72 [0.59-0.87]*** |
|  | **(O) ECOG 2 pre-trial effect** | **216 (ECOG 2 RCT: 101, ECOG 2 RWD: 169; ECOG 3 RWD: 47 )** | **ECOG 2 RCT: (ref), ECOG 2 RWD: 0.79 [0.48-1.3] , ECOG 3 RWD: 1.26 [0.74, 2.1]** |
|  | (O) Including censored records for study futility | 772 (544, 228) | 0.67 [0.56-0.81]*** |
|  | (O) Removing all censored patients | 433 (210, 223) | 1.5 [1.2-1.8]*** |
|  | (O) Propensity score including also the censoring as variable | 456 (228, 228) | 1.5 [1.2-1.8]*** |
|  | (O) Oversampling RWD simulating RCT censoring | 922 (461, 461) | 1.5 [1.2-1.8]*** |

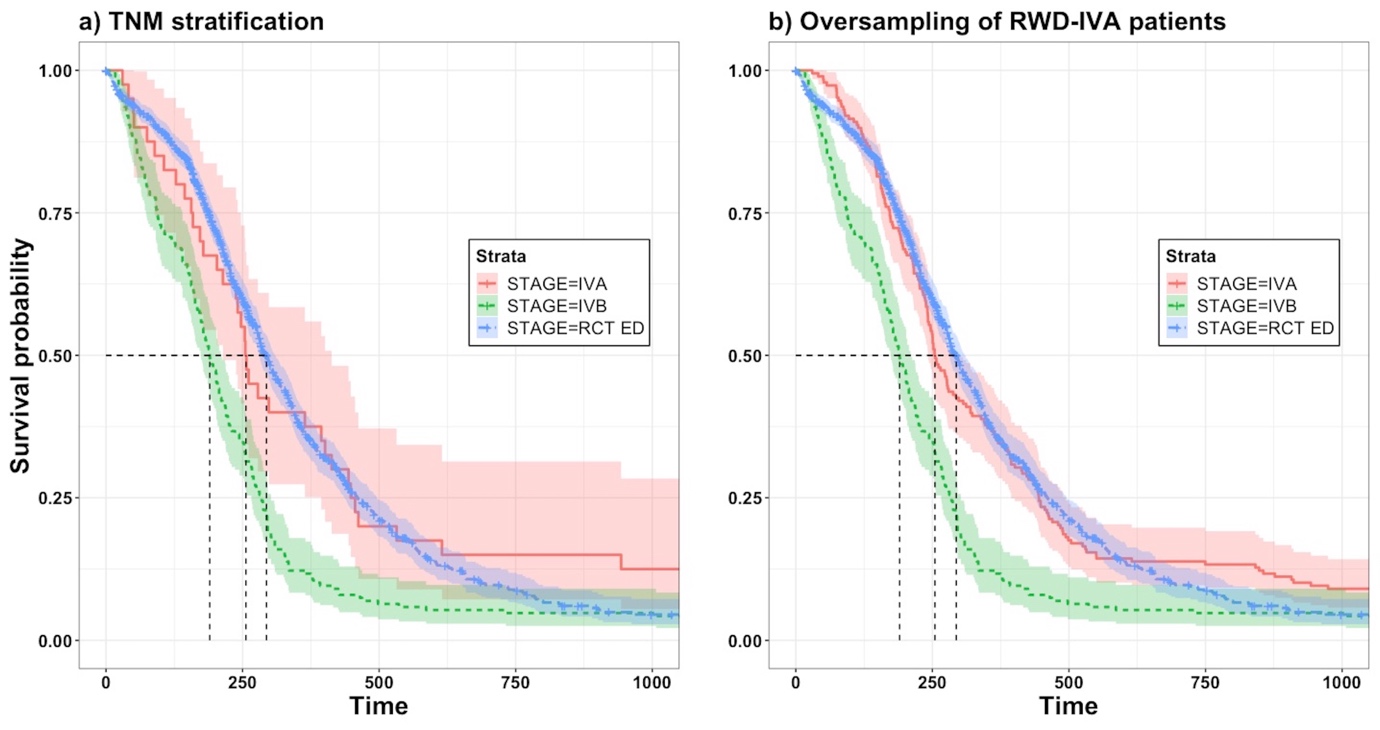

Figure S2. Overall Kaplan Meier curves of STAGE sub-stratification: a) TNM stratification (n=1224), b) TNM oversampling (n=1372).

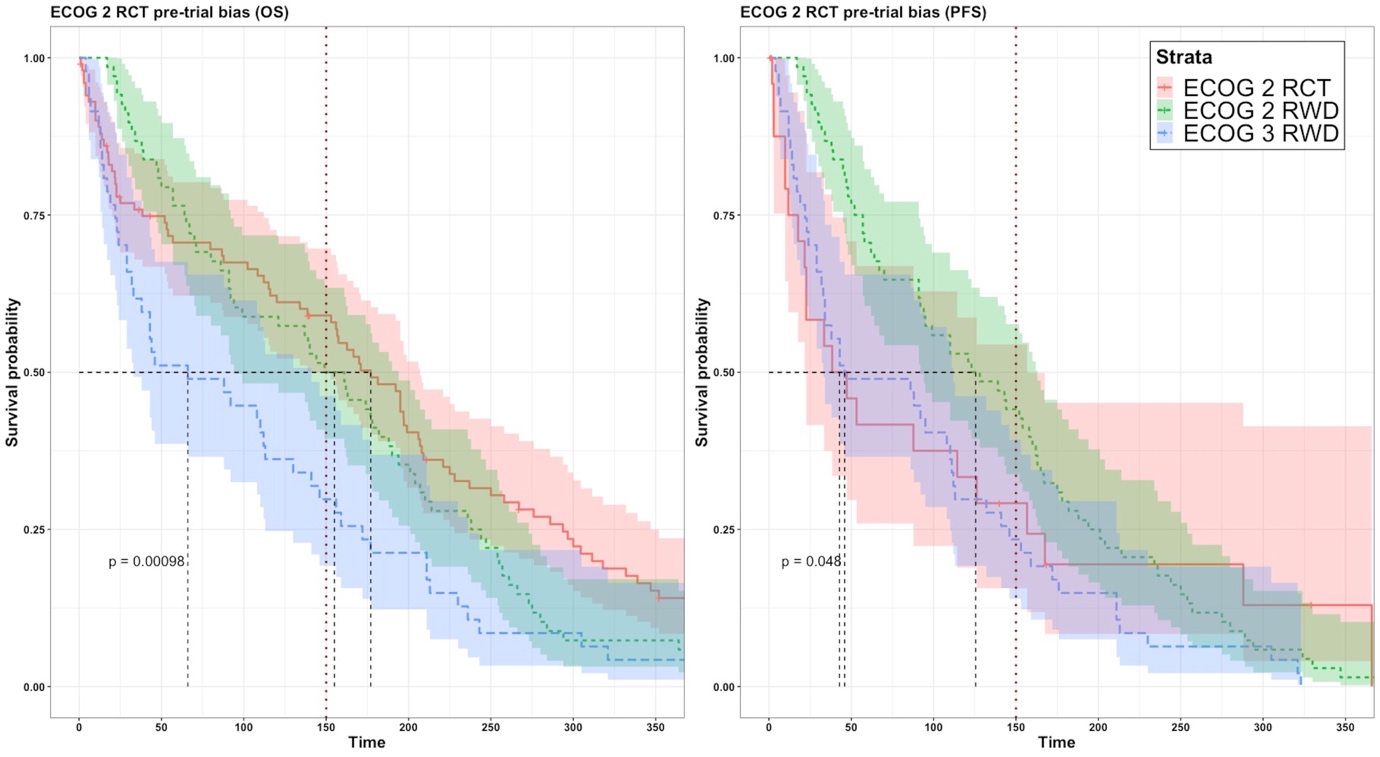

Figure S3. ECOG 2 pre-trial bias. Trial patients with ECOG 2 is compared with real-world ECOG 2-3 patients. Trial patients presents more similarities with real-world ECOG 3 patients.

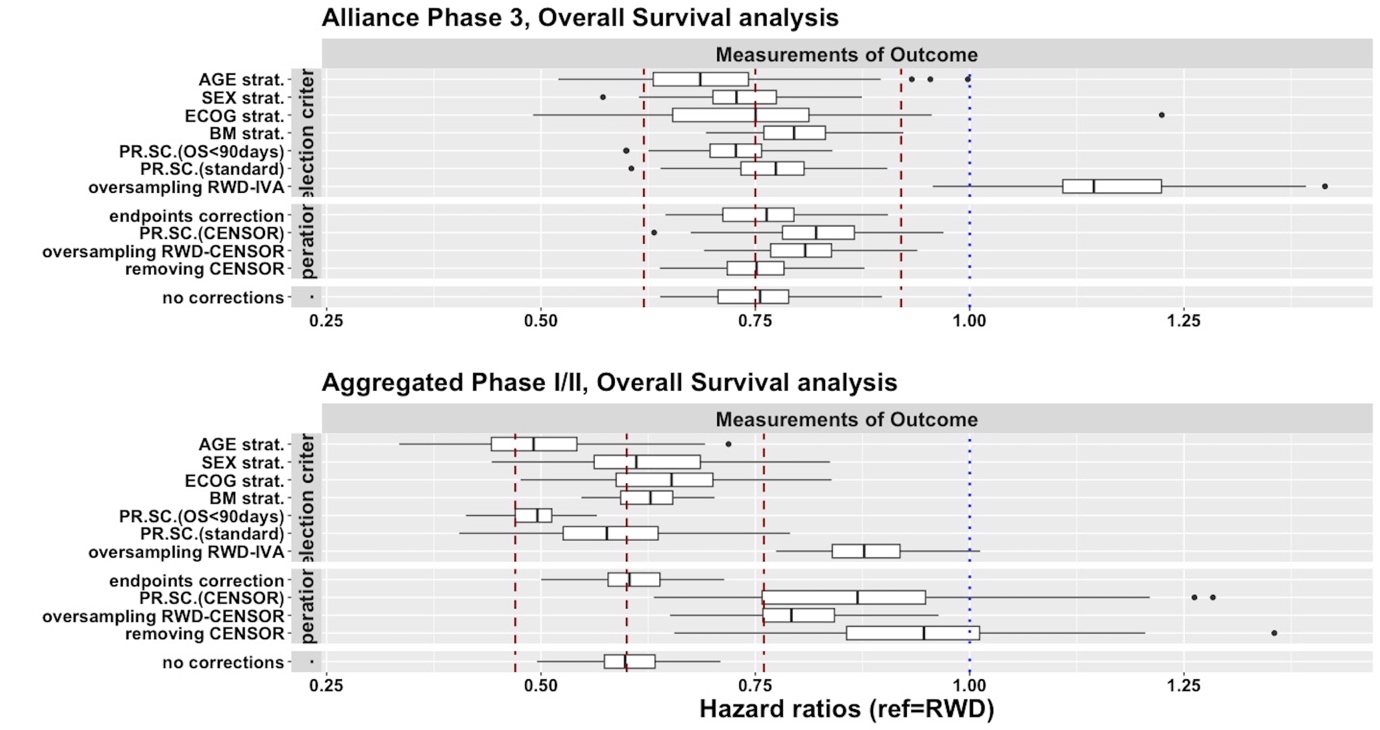

Figure S4. Matching Simulation results for overall survival hazard ratios for PDS_Allianc and the aggregated cohort of Phase Ib-II studies. Strat.: stratification, BM: brain metastases, PR.SC.: propensity score, RWD: real-world data. Hazard ratio and 95% confidence level interval with the whole cohort is reported in red dotted lines.

1. RCT: random clinical trial, RWD: real world data. BM: Brain Metastasis, OS: overall survival, PFS: progress free survival
2. Bold text: detected similarity of outcomes, Underlined text: detected switch of survival outcome (better prognosis for real-world patients). (S): selection criteria, (O): operations, MO: measurement of outcomes, RCT: randomize clinical trials, RWD: real-world data. For the hazard ratios is reported the 95% confidence level interval. (*) p-value<0.05, (**) p<0.01, (***) p<0.001.
